# Evaluating and Improving the Performance and Racial Fairness of Algorithms for GFR Estimation

**DOI:** 10.1101/2024.01.07.24300943

**Authors:** Linying Zhang, Lauren R. Richter, Tevin Kim, George Hripcsak

**Affiliations:** Department of Biomedical Informatics, Columbia University, New York, NY, USA; Institute for Informatics, Data Science, and Biostatistics, Washington University in St. Louis St. Louis, MO, USA

**Keywords:** algorithmic fairness, glomerular filtration rate, predictive modeling, machine learning, electronic health record

## Abstract

Data-driven clinical prediction algorithms are used widely by clinicians. Understanding what factors can impact the performance and fairness of data-driven algorithms is an important step towards achieving equitable healthcare. To investigate the impact of modeling choices on the algorithmic performance and fairness, we make use of a case study to build a prediction algorithm for estimating glomerular filtration rate (GFR) based on the patient’s electronic health record (EHR). We compare three distinct approaches for estimating GFR: CKD-EPI equations, epidemiological models, and EHR-based models. For epidemiological models and EHR-based models, four machine learning models of varying computational complexity (i.e., linear regression, support vector machine, random forest regression, and neural network) were compared. Performance metrics included root mean squared error (RMSE), median difference, and the proportion of GFR estimates within 30% of the measured GFR value (P30). Differential performance between non-African American and African American group was used to assess algorithmic fairness with respect to race. Our study showed that the variable race had a negligible effect on error, accuracy, and differential performance. Furthermore, including more relevant clinical features (e.g., common comorbidities of chronic kidney disease) and using more complex machine learning models, namely random forest regression, significantly lowered the estimation error of GFR. However, the difference in performance between African American and non-African American patients did not decrease, where the estimation error for African American patients remained consistently higher than non-African American patients, indicating that more objective patient characteristics should be discovered and included to improve algorithm performance.

## I. Introduction

Amid calls for greater racial equity throughout society, medicine too must adapt to alleviate racial biases and predispositions with a “race-conscious” approach [1]. While human experts remain the ultimate decision-makers in most clinical settings, clinical algorithms play an increasingly important role in assisting clinicians in the process of decision-making across a wide variety of medical specialties. Many of the existing algorithms use race as a predictor to predict patient outcomes, some of which have been shown to underestimate the health risk of minority patients, limiting access to timely and specialized care [2]. An example of such an algorithm is the original serum creatinine-based Chronic Kidney Disease Epidemiology Collaboration (CKD-EPI_cr_) equation [3]. Developed in 2009, it uses serum creatinine levels and demographic modifiers, including one that isolates patients identified as Black, to calculate the estimated glomerular filtration rate (eGFR).

Glomerular filtration rate (GFR) is a measure of renal function that is readily used to influence clinical decisions surrounding kidney disease [4]. The most accurate measures of GFR are taken with exogenous markers (*e*.*g*., inulin, iothalamate, and iohexol) over a 24-hour period, but their clinical use is severely limited due to impracticality [5], [6]. As a result, eGFR (as calculated by the serum creatinine-based CKD-EPI_cr_ equation) is the most common clinically used method of determining GFR followed by the urinary clearance of creatinine over 24-hours. Recent studies have also suggested that the use of serum cystatin C levels could increase eGFR accuracy, but this practice is not widespread [7], [8]. While eGFR is not the only factor influencing the clinical decision-making process, eGFR thresholds are often used to assist in that regard. For example, guidelines suggest patients with eGFR values under 30mL/min/1.73m^2^ should be referred to nephrology specialists, while patients with eGFR values under 20mL/min/1.73m^2^ should be considered for kidney transplantation; these same guidelines also dictate how the prescription of nephrotoxic and renally excreted medications, many unrelated to nephrology, requires consideration of eGFR [9], [10].

As such, given the clinical importance of eGFR in the diagnosis and treatment of chronic kidney disease (CKD), discussions around the ethics of including a race modifier in estimating GFR have been around since the introduction of the CKD-EPI_cr_ equation [1]–[3], [7]–[9], [11]–[14]. Due to the race modifier in the CKD-EPI_cr_ equation, patients who identify as Black are estimated to have 16% higher GFR than their non-Black counterparts. Moreover, the inclusion of race in the original CKD-EPI_cr_ equation lacks biologically substantiated evidence [11], [12], and removing race from the equation could have a significant impact on recommended care for Black patients (*e*.*g*., increasing CKD diagnoses among Black adults could improve access to specialist care and kidney transplantation).

Partially in response to the racial and ethical concerns surrounding the 2009 CKD-EPI equation, in 2021, a refit serum creatinine-based Chronic Kidney Disease Epidemiology Collaboration (CKD-EPI_cr_Refit_) equation [7] was developed without the consideration of race and was recommended for national use by a combined National Kidney Foundation and American Society of Nephrology (NKF-ASN) task force [8]. In contrast to the current equation, which adversely affects Black patients because of overestimation of GFR, the new equation without race correction has statistical bias more evenly distributed between the two racial groups: it underestimates GFR for Black patients and overestimates GFR for non-Black patients [7]–[9], [11], [14]. Until the new equation receives a wider adoption in clinical practices, it is unclear whether the race-free equation would actually impact care for patients across all racial groups and its impact on alleviating health disparities between Black and non-Black patients.

It is important to understand the legal and ethical context when developing and deploying algorithms in healthcare, but such assessments are unlikely to be sufficient without rigorous statistical quantification of algorithmic performance and fairness. In the past five years, the statistics and machine learning community has explored extensively different ways of quantifying fairness of algorithms, such as statistical parity, equalized odds, and equal opportunity [15]. These fairness definitions have been studied in various social contexts, such as college admission, criminal justice, and employment, but their application to clinical algorithms remains to be better understood. In some of the recent studies, challenges with applying these definitions have been reported, with the major challenge being that these definitions cannot be simultaneously satisfied, and how to choose among these fairness definitions remains an open question [16].

In this study, we investigate what factors impact the performance of fairness of algorithms for GFR estimation. We use an error-based fairness notion to statistically quantify the deviation from fairness of an eGFR model. We compare the performance and fairness of models under different modeling designs, including the choice of including or excluding race in the equation, the amount of input information, and the complexity of machine learning models.

## II. METHODS

### A. Cohort Definition and Extraction

The cohort for this study was built with de-identified data from Columbia University Irving Medical Center’s (CUIMC) electronic health records (EHR) collected from 1980 to 2021. The entry event for our cohort was defined as the measurement of urine creatinine over a 24-hour period (Ucr). Members of the cohort were required to have all prerequisite data for eGFR and urine creatinine-based measured GFR (mGFRcr) calculation. These included age, gender, race, serum creatinine levels (Scr), and body surface area (BSA), which could alternatively be calculated with weight and height as per the Du Bois formula [17]. In addition to the data points mentioned above, diagnosis and medication data, from the 365-day period preceding Ucr measurement, were extracted for each participant.

In an effort to minimize aberrations and extraneous cases, patients with documented pregnancies within a month of creatinine measurements were excluded, and only those between the ages of 18 and 100 were included. Furthermore, due to a lack of data, only patients who identified as “Black or African-American”, “Asian”, or “White” were incorporated into this study.

The patients’ true GFR was approximated with 24-hour urinary creatinine clearance, denoted as mGFRcr, and calculated as follows [5], [18], [19]:

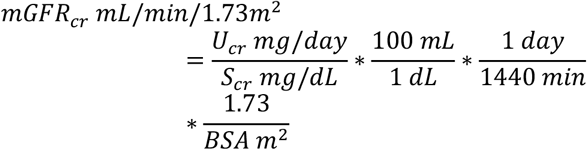

### B. Development of Models for GFR Estimation

In this study, we compared three distinct approaches for estimating GFR: 1) CKD-EPI equations, 2) epidemiological models, and 3) EHR-based models, in the order of increasing number of input features. In terms of model complexity, the CKD-EPI equation is a linear regression on the logarithmic scale of GFR with two splines for creatinine. The epidemiological models and EHR-based models go beyond linear and parametric models to include nonlinear and nonparametric models. All models were trained with and without the variable race.

The CKD-EPI equations were used as a baseline representative of current medical practice. The CKD-EPI equation is a linear regression on the logarithmic scale of the measured GFR with age, sex, two splines for creatinine, and in some cases, race. To ensure fair comparison between CKD-EPI equations and other modeling approaches, we did not use the parameters from the existing CKD-EPI equations, but instead, trained a CKD-EPI equation on our own database (CUIMC CKD-EPI). The CUIMC CKD-EPI equation had the same functional form of the 2009 CKD-EPI and the 2021 CKD-EPI equations, and differed only in the value of the parameters.

In addition to the variables used by the CKD-EPI equation, the epidemiological models included four additional variables: hypertension, type 2 diabetes mellitus, height, and weight. These features were found to be associated with glomerular filtration rate and/or kidney function by other published studies [20]. The EHR-based models included the full medical history of patients (e.g., diagnoses, medications, labs ordered, vital signs measured) within one year prior to the cohort entry.

Because both epidemiological models and EHR-based models include a large-set of covariates, we fit models of varying complexity–linear regression (LR), random forest (RF), support vector machine (SVM), and neural network (NN)–to best leverage the large set of predictors. For model training, we bootstrapped the data and split the bootstrapped dataset into an 80% training set and a 20% testing set. The model was trained and hyperparameters were selected based on 5-fold cross-validation on the 80% training set. Then, the model was applied to the 20% test set to estimate GFR. Performance metrics (described below) were computed on the test set. We randomly bootstrapped 30 datasets and reported the mean and 95% confidence interval of all performance metrics across the 30 bootstrapped datasets.

### C. Evaluation Metrics of Algorithmic Performance

Three evaluation metrics were used throughout this study: root mean square error (RMSE), median difference, and the proportion of GFR estimates within 30% of the measured GFR value (P30). These metrics are commonly reported in other GFR estimation studies to compare model performance [3], [17], [21].

The RMSE measures the average magnitude of the errors, without considering their direction. It ranges from zero to infinity and lower value means better performance. The median difference was calculated as the median mGFR minus the median eGFR value. In contrast to RMSE, median difference does not measure the average but the median of the error distribution. It ranges from negative to positive infinity with a positive value indicating underestimation of the GFR.

The P30 accuracy was used in part to put a greater emphasis on accuracy at lower ranges of GFR as the accuracy of GFR estimation is much more clinically important when a patient’s kidney function is severely reduced (i.e., the true GFR is low). P30 accomplishes this as the 30% range needed for an estimate to be considered accurate shrinks as the measured GFR decreases. Higher P30 means better estimation, and a P30 value of 80-90% is considered to be acceptable and above 90% is preferred for GFR evaluation in many circumstances [7].

### D. Definition and Evaluation of Algorithmic Fairness

An increasingly adopted approach to assess fairness of predictive algorithms is to compare the distribution of error across sensitive groups (e.g., racial groups). However, previous studies have shown that an algorithm can be fair by some error metrics but not others [15], [22]. This is more extensively studied for predicting binary outcomes (e.g., mortality), and less for continuous outcomes, like in this case, GFR.

We adopt this equal-error approach to assess fairness of algorithms for GFR estimation. We computed all three metrics for African American and non-African American separately, in addition to calculating these metrics for the entire study cohort. A delta value (Δ) at zero means the model has equal performance between race groups, and a positive value means that the model has higher error for African American patients.

## III. RESULTS

### A. Cohort Characterization

The study cohort consists of 2352 non-pregnant adult patients, of whom 610 (25.9%) are documented as African-American, and 1742 (74.1%) are documented as White or Asian. Selected characteristics of the cohort are shown in Table 1. African American patients were on average 6 years younger (mean [SD], 54[16.0] years old) compared to Non-African American patients (mean [SD], 60[15.6] years old). The mean of measured urine creatinine-based GFR (mGFRcr) of African American patients was 6 mL/min/1.73m2 lower (mean [SD], 63[48.9]) compared to non-African American patients (mean [SD], 69[44.5]), indicating worse kidney function among African American patients; the standard deviation of mGFRcr was 4.5 mL/min/1.73m2 higher for African American patients, indicating more heterogeneity in this racial group. Moreover, more African American patients had hypertension (African American: 64.4% vs non-African American: 46.2%), type 2 diabetes (African American: 30.5% vs non-African American: 18.8%), and chronic kidney disease (African American: 32.5% vs non-African American: 21.8%), and as a result, anti-hypertensive (e.g., heparin, amlodipine) and antiglycemic medications (e.g., glucagon, insulin aspart) were more prevalent among African American patients.

**TABLE 1.** Cohort characterization.

| Characteristic | Total (N=2352) | African American (N=610) | Non-African American (N=1742) |
| --- | --- | --- | --- |
| **Demographics** |  |  |  |
| Gender, No. (%) | 1157(49.2) | 319(52.3) | 838(48.1) |
| Age, Mean (SD) | 58(15.9) | 54(16.0) | 60(15.6) |
| Measurement, Mean (SD) |  |  |  |
| mGFRcr (mL/min/1.73m<sup>2</sup>), Mean (SD) | 68(45.7) | 63(48.9) | 69(44.5) |
| Serum Creatinine (mg/dL) | 1(1.6) | 2(2.1) | 1(1.4) |
| Weight (kg) | 77(20.5) | 83(21.3) | 75(19.9) |
| Height (cm) | 168(10.2) | 169(10) | 168(10.3) |
| **Diagnosis, No. (%)** |  |  |  |
| Essential hypertension | 1197(50.9) | 393(64.4) | 804(46.2) |
| Chest pain | 594(25.3) | 216(35.4) | 378(21.7) |
| Type 2 diabetes mellitus | 514(21.9) | 186(30.5) | 328(18.8) |
| Type 2 diabetes mellitus without complication | 448(19.0) | 166(27.2) | 282(16.2) |
| Chronic kidney disease | 578(24.6) | 198(32.5) | 380(21.8) |
| Acute renal failure syndrome | 791(33.6) | 250(41.0) | 541(31.1) |
| Dyspnea | 943(40.1) | 285(46.7) | 658(37.8) |
| Obesity | 236(10.0) | 97(15.9) | 139(8.0) |
| Iron deficiency anemia | 145(6.2) | 71(11.6) | 74(4.2) |
| Chronic kidney disease due to hypertension | 280(11.9) | 106(17.4) | 174(10.0) |
| **Medication Ingredient, No. (%)** |  |  |  |
| Heparin | 973(41.4) | 311(51.0) | 662(38.0) |
| Amlodipine | 641(27.3) | 220(36.1) | 421(24.2) |
| Glucose | 1196(50.9) | 361(59.2) | 835(47.9) |
| Carvedilol | 289(12.3) | 124(20.3) | 165(9.5) |
| Hydralazine | 283(12.0) | 120(19.7) | 163(9.4) |
| Glucagon | 469(19.9) | 163(26.7) | 306(17.6) |
| Labetalol | 172(7.3) | 85(13.9) | 87(5) |
| Lisinopril | 316(13.4) | 122(20) | 194(11.1) |
| Insulin aspart, human | 413(17.6) | 147(24.1) | 266(15.3) |
| Acetaminophen | 1535(65.3) | 436(71.5) | 1099(63.1) |

### B. Performance and Fairness Evaluation

The performance and racial fairness of three modeling approaches: CUIMC CKD-EPI equations, epidemiological models with random forest (EPI-RF), and EHR-based models with random forest (EHR-RF) for GFR estimation were shown in Figure 2.

**Fig. 1.**
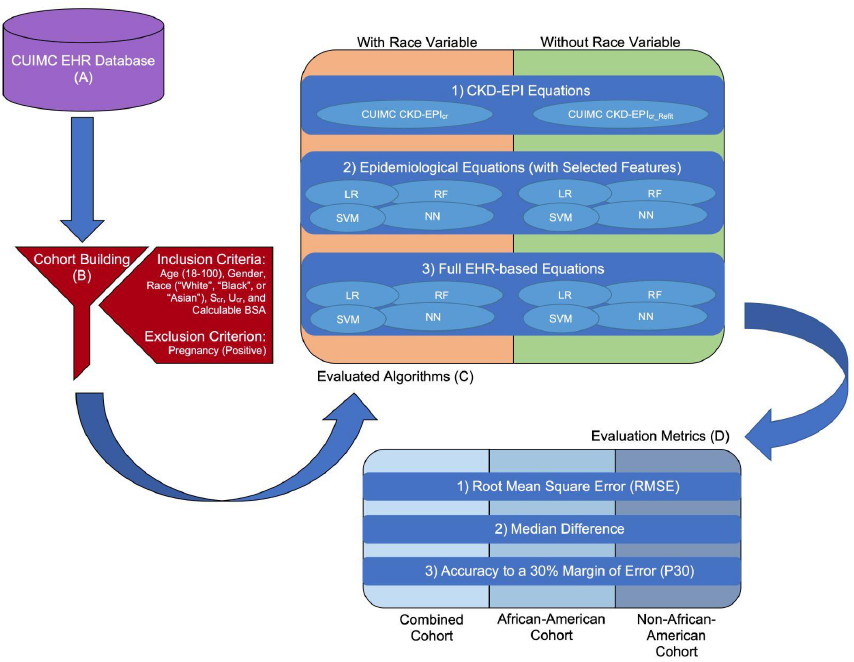
An overview of the methodology for this study: (A) Data for this study originated from Columbia University Irving Medical Center’s electronic health record database. (B) The cohort was selected based on listed inclusion and exclusion criteria after which all relevant patient data were extracted from the database. A criterion without parentheses simply refers to a non-null value, while criteria with parentheses must have one of the values indicated. (C) Each ellipse represents an algorithm evaluated over the course of this study, and they are categorized based on the amount of data considered (in increasing order from 1 to 3) and the presence of the race variable. CKD-EPI equations are representative of current medical practice, while epidemiological and EHR-based equations were developed during this study as linear regression, random forest, support vector machine, and artificial neural network models. Furthermore, the adjusted CKD-EPI equations are equivalent to the CKD-EPIcr equation with the exception of refit coefficients. (D) Each of the three evaluation metrics was calculated for each algorithm from part C, and calculations were done separately for each cohort.

**Fig. 2.**
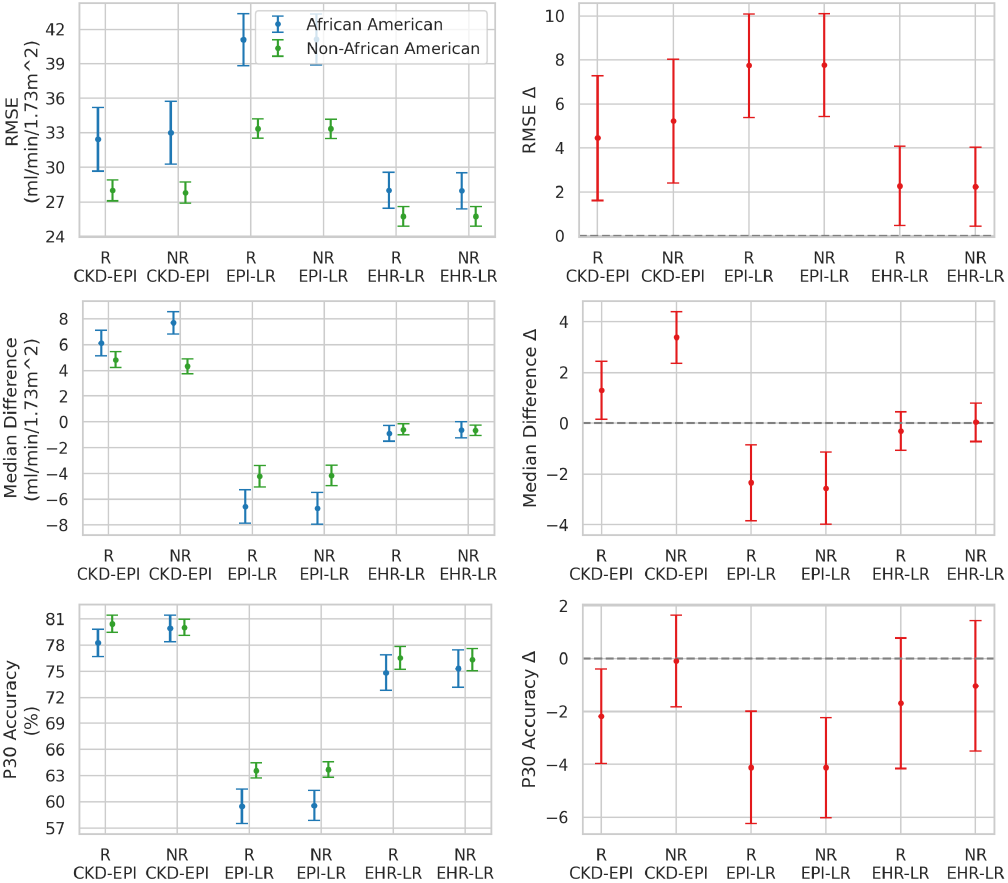
Performance and racial fairness of random forest models for GFR estimation. Three modeling approaches were compared in the study, CUIMC CKD-EPI, EPI-RF, and EHR-RF models. Models were trained with race (R) and without race (NR). Performance was evaluated with root mean square error (RMSE), median difference, and accuracy within 30% of the measured value (P30). Racial fairness was quantified by the difference in performance between African American and non-African American groups, denoted by delta (Δ), and delta equals zero indicating fairness. while the error bar represents a 95% confidence interval.

First, we report findings regarding the impact of the variable race on GFR estimation. We begin with the current clinical algorithm, CKD-EPI equation. Comparing the two versions (with and without race) of CUIMC CKD-EPI equations, we found that removing race affected the median difference for both racial groups, but no significant impact on other performance metrics. Specifically, the median difference for African American patients increased after race was removed, and decreased for non-African American patients. This change in median difference due to the removal of race was also observed in Delgado et al [8]. The impact of race on RMSE and P30 was not significant. Based on the comparison of CKD-EPI with and without race as a predictor, it was unclear which approach was better because the performance did not significantly improve.

Moving beyond CKD-EPI equations, we found that the impact of race diminished and became almost negligible across all performance metrics as more variables that are potentially informative about GFR were included in the prediction. This finding implies that the association between race and GFR could be explained by other predictors, and more objective predictors, such as diseases and physiological measurements, should be identified to better estimate GFR.

Second, including more predictors resulted in lower estimation error of GFR estimation for both racial groups. As an example, we compared the three modeling approaches that did not include race (the inclusion of race had negligible impact on the performance). The RMSE for African Americans decreased from 33.0 (95% CI: [30.2, 35.9]) in the CKD-EPI model to 24.5 (95% CI: [22.3, 26.5]) in the EPI-RF model and to 23.5 (95% CI: [21.6, 25.9]) in the EHR-RF model. The RMSE for non-African Americans also decreased from 27.8 (95% CI: [27.0, 28.8]) in the CKD-EPI model to 21.3 (95% CI: [20.5, 22.0]) in the EPI-RF model and to 19.4 (95% CI: [18.7, 20.0]) in the EHR-RF model. Improvement was observed on the other two evaluation metrics. The median difference for African American patients decreased from 7.82 (95% CI, [6.99, 8.65]) to less than one in the epidemiological-RF and EHR-RF models. Similarly, the median difference for non-African American patients decreased from 4.32 (95% CI, [3.82, 4.82]) to near zero in the epidemiological-RF and EHR-RF models. The accuracy P30 slightly improved from about 80% for both race groups in the CKD-EPI model to about 81% for both race groups in the EHR-RF models, but this change was not significant. These findings suggest that the additional epidemiological factors included and the increased model complexity can improve the estimation of GFR.

Third, including more predictors did not lead to more equal distribution of error between African American and non-African American patients. The RMSE for African American patients was consistently higher than non-African American patients by about 4mL/min/1.73m^2^ across all models tested. Similarly, the P30 accuracy was lower for African American patients than non-African American patients by about 1.5%. Including more predictors led to better performance (lower error) for both racial groups, but did not lead to the error distributed more evenly between the two racial groups. This could be due to the lower sample size of African American patients in the study cohort.

Performance of other machine learning models are shown in Fig. 3-5. The trend regarding performance and fairness does not vary significantly across non-linear machine learning models (RF, SVM, and NN) tested in this study.

**Fig. 3.**
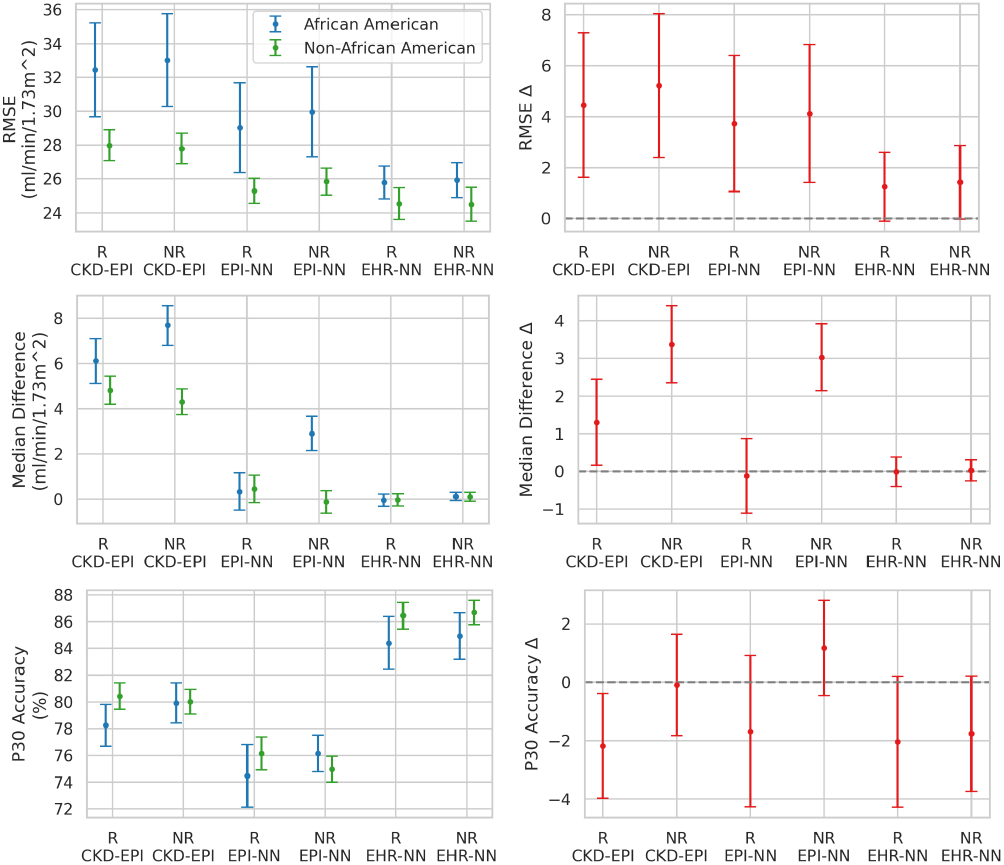
Performance and racial fairness of linear regression models for GFR estimation.

**Fig. 4.**
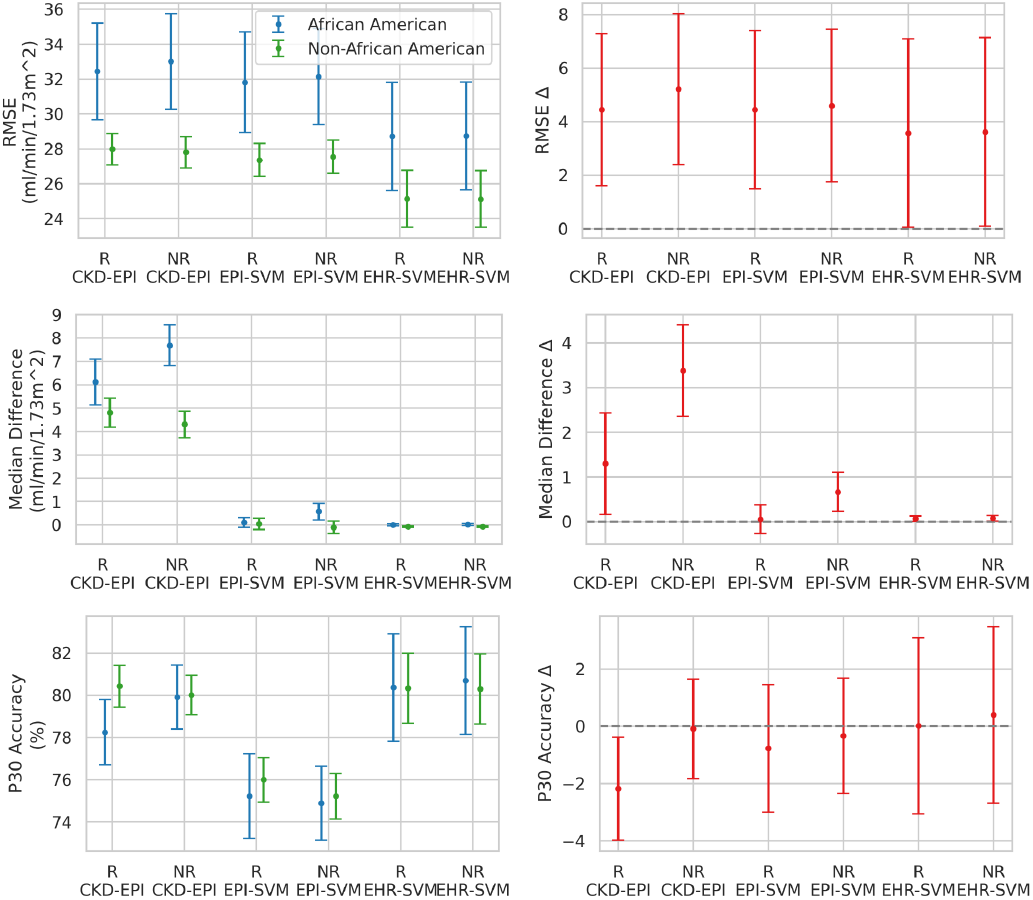
Performance and racial fairness of support vector machine (SVM) models for GFR estimation.

**Fig. 5.**
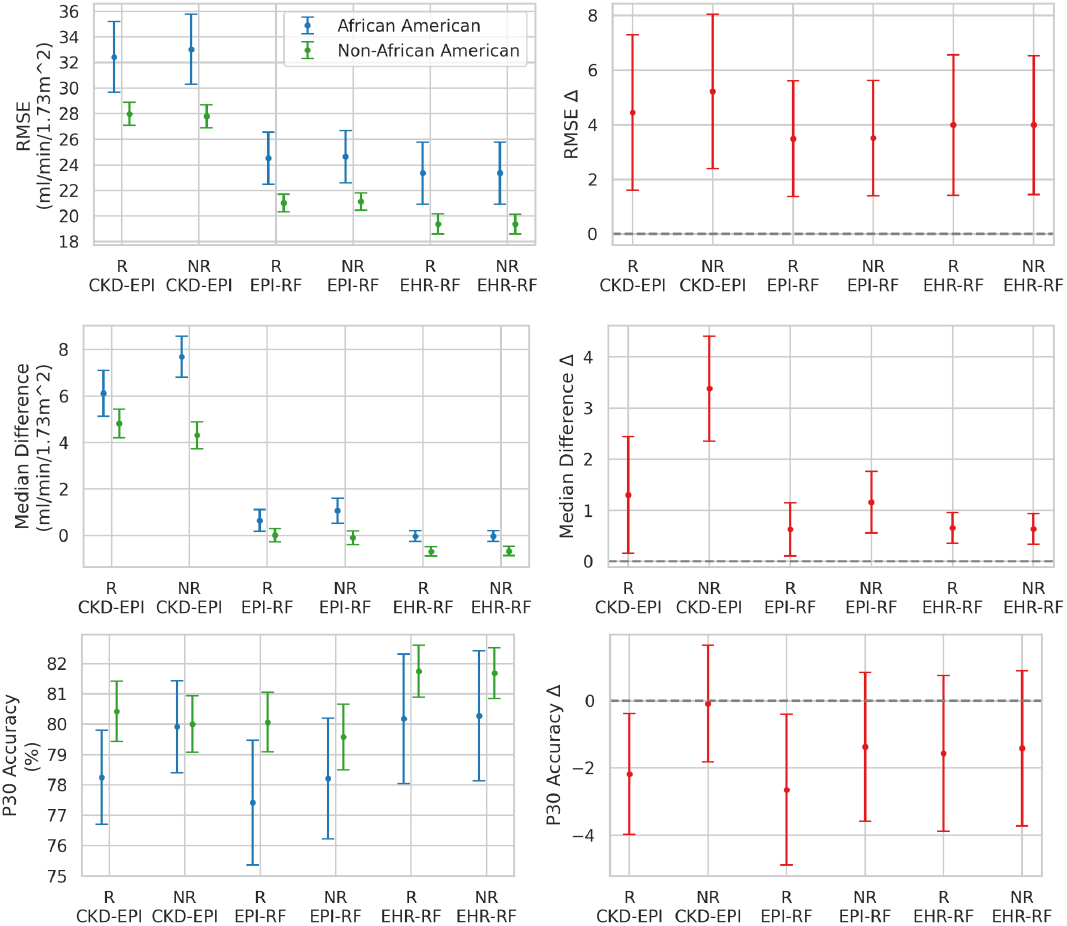
Performance and racial fairness of neural network (NN) models for GFR estimation.

## IV. DISCUSSION

As more data-driven algorithms are being used in clinical settings to assist clinical decision-making, it is important to understand what factors can influence the performance and fairness of such algorithms, and how to mitigate bias in developing clinical algorithms to ensure fair treatment of all individuals.

The National Kidney Foundation and American Society of Nephrology (NKF-ASN) task force reached a consensus that estimating equations that do not incorporate race are desirable and needed [8]. We agree on this move given the negligible effect of race on improving GFR estimation in this study. Furthermore, we suggest to seek more objective features that are predictive of the outcome of interest. One potential reason that race appears in many clinical algorithms in the first place is that race is associated with many other health-related factors, including genetic, social, environmental, and behavioral factors. If some of these health-related factors are more heterogeneous within certain racial groups, then the heterogeneity can lead to more variability in the outcome we try to predict in certain racial groups. Rather than including race, a bad proxy for various determinants of health, we should devote our effort to identifying real causes of the outcome we try to estimate. In this study, we show that the inclusion of common comorbidities, height, and weight can dramatically decrease estimation error. The inclusion of all conditions and medications from the medical record may further decrease the estimation error.

Besides including the causal factors in the algorithm, modeling choices can also significantly impact model performance and fairness. Accurate estimation of GFR involves multiple interacting factors in a complex nonlinear relationship. Machine learning models are known to be powerful in modeling complex data relationships. Both the original and the 2021 refit CKD-EPI equations were built on least-squares linear regression models [3], [7], while machine learning models have been demonstrated to be powerful for various clinical applications, but their strength in GFR estimation has been underexplored. One study explored ensemble models and another study explored deep learning models for GFR estimation [21], [23]; however, these do not thoroughly evaluate their results in the context of reducing racial biases. Our study shows that nonlinear machine learning models improve the estimation accuracy for both Black and non-Black patients, but the gap in performance between these two groups does not change significantly.

While this work focuses on a particular equation in nephrology, the findings and questions that remained are likely to be encountered in other medical disciplines where race-adjusted clinical algorithms exist. Together, we should move away from using race in clinical equations and instead, seek more objective features that are predictive of the outcome of interest. Furthermore, race-free clinical algorithms are not the final goal and may very well be the beginning. As shown by our study, race-free models can still have differential performance across racial groups, with African Americans experiencing higher estimation error than non-African Americans. Future algorithm development should consider recruiting more patients from the minority and historically disadvantaged groups, as this could improve the sample imbalance problem. Last but not least, errors can be quantified in many ways, and there is no consensus on what statistical metrics are better than others. We show that RMSE, median difference, and P30 could be useful in assessing algorithms for GFR estimation, but what metrics to use is likely context-dependent and should be considered carefully in future studies.

This work has several limitations. First, categorization of race into two groups does not adequately represent the diversity within and among race groups in the United States. The choice of binary race is partially due to the lack of Hispanic ethnicity, Native American, and other racial minorities in the cohort. Further research should consider leveraging multiple data sources to include patients from diverse racial and ethnic backgrounds. Second, only about 3000 patients’ EHR data from Columbia University Irving Medical Center were used throughout the course of this study. As a result, the results might not be generalizable to other databases. Lastly, the measured GFR is derived based on 24-hour urine clearance of creatinine. Some studies showed that creatinine clearance overestimates true GFR due to tubular creatinine secretion [5]. However, we do not expect our conclusions about algorithmic fairness to change because there is no evidence suggesting that this measurement error is differentially distributed across racial groups.

## V. Conclusion

This study is reflective of the greater need to reevaluate the historical use of race in medicine, especially as the medical community accepts race as a “social, not biological, construct” [24]. As such, it is of the utmost importance that we find alternatives to using race in clinical algorithms and decision-making in order to further improve algorithmic performance and reduce differential impact of the algorithm across racial groups. Hopefully, the broader use of EHR data can account for variations previously associated with race. This study shows that the use of more advanced machine learning coupled with the leveraging of more patient data can have a positive effect on GFR estimation regardless of the patient’s racial background by increasing overall performance.

## Data Availability

The data that support the findings of this study are available on request from the corresponding author. The data are not publicly available due to privacy or ethical restrictions.

## Acknowledgment

This work was supported by National Institutes of Health (NIH) R01LM006910.

## Footnotes

Identify applicable funding agency here. If none, delete this text box.

